# Modeling of the COVID-19 Cases in Gulf Cooperation Council (GCC) countries using ARIMA and MA-ARIMA models

**DOI:** 10.1101/2021.05.27.21257916

**Authors:** Rahamtalla Yagoub, Hussein Eledum

## Abstract

Coronavirus disease 2019 (COVID-19) is still a great pandemic presently spreading all around the world. In Gulf Cooperation Council (GCC) countries, there were 1015269 COVID-19 confirmed cases, 969424 recovery cases, and 9328 deaths as of 30^th^ Nov. 2020. This paper, therefore, subjected the daily reported COVID-19 cases of these three variables to some statistical models including classical ARIMA, k^th^ SMA-ARIMA, k^th^ WMA-ARIMA, and k^th^ EWMA-ARIMA to study the trend and to provide the long-term forecasting of the confirmed, recovery, and death cases of the novel COVID-19 pandemic in the GCC countries. The data analyzed in this study covered the period starting from the first case of coronavirus reported in each GCC country to Nov 30, 2020. To compute the best parameter estimates, each model was fitted for 90% of the available data in each country, which is called the in-sample forecast or training data, and the remaining 10% was used for the out-of-sample forecast or testing model. The AIC was applied to the training data as a criterion method to select the best model. Furthermore, the statistical measure RMSE was utilized for testing data, and the model with the minimum AIC and minimum RMSE was selected. The main finding, in general, is that the two models WMA-ARIMA and EWMA-ARIMA, besides the cubic linear regression model have given better results for in-sample and out-of-sample forecasts than the classical ARIMA models in fitting the confirmed and recovery cases while the death cases haven’t specific models.

## 1. Introduction

Coronavirus disease 2019 (COVID-19) is an infectious disease caused by severe acute respiratory syndrome coronavirus 2 (SARS-CoV-2). The first case was registered in Wuhan, China, in December 2019. It has since spread worldwide, leading to an ongoing pandemic. Most of the people around the world have been infected with the new COVID-19 virus nowadays; the people of the Gulf Cooperation Council (GCC) countries were no exception to that. GCC countries adopted many severe strategies and policies to face the new crisis, such as early diagnosis, isolated infected people, and social distancing. The first COVID-19 confirmed in the GCC area was reported in UAE on Jan 29, 2020, and flowed by Bahrain on Feb 21, 2020, Kuwait, and Oman on Feb 24, 2020, Qatar on Feb 27, 2020, and KSA on Mar 2, 2020. Confirmed, recovery, and death cases are recorded daily in each of these countries. Accordingly, the ultimate requirement is to create an efficient statistical methodology that can efficiently predict morbidity cases. Thus, it can aid in decision-making and logistical planning in healthcare systems for coming challenges. The statistical forecast models are always conducted to predict the disease’s behavior in the future, and in this way, it may decrease and restrain in the pandemic.

Recently, many studies of COVID-19 have been conducted in the GCC countries; some of these studies used measures of descriptive statistics to explain the disease prevalence, incidence, and underline prevention and control techniques applied in these countries; while others used mathematical models to analyze and predict the evolution of the disease. Alandijany et al. (2020) reviewed the status of COVID-19 in GCC countries, summarized the control measures taken by each government, and highlighted some future challenges. They recommended that these countries must take severe appropriate precautions to limit the spread of infection. Sharif et al. (2020) utilized the SIRD and smoothing spline regression models to predict the number of cases in the Eastern Mediterranean Region including Saudi Arabia, Iran, and Pakistan. They concluded that the cumulative infected cases were expected to grow exponentially during the study period from Jan 29 till Apr 14, 2020. Zuo et al. (2020) provided a brief comparison of the COVID-19 events, which involve the total confirmed cases, total deaths, total recovery, and active cases that have been reported in the Asian countries up to Apr 8, 2020. Moreover, they introduced a new family of statistical models and proposed a particular submodel called flexible extended Weibull distribution. Abuhasel et al. (2020) applied the classical SIR model besides the ARIMA model to forecast the prevalence and recovery rates of the COVID-19 pandemic; the two models were applied to the daily data from Mar 3 to Jun 30, 2020. Ayinde et al. (2020) used the statistical curve model to model the daily cumulative confirmed, discharged, and death cases in Nigeria for the period beginning from Feb 27, 2020, until Apr 30, 2020. They concluded that the Cubic Linear Regression with AR (1) models are the best ones. Elhassan and Gaafar (2020) employed ARIMA and logistic growth models to predict the cumulative confirmed, recovery, and death cases of COVID-19 in Saudi Arabia between 2^nd^ March 2, 2000, and Jun 21, 2020. They inferred that ARIMA(0,2,0), ARIMA(1,2,0), and ARIMA(1,2,3) were the most useful models to fit the cumulative confirmed, recovery, and death cases, respectively. Singh et al. (2020) developed the ARIMA to model the daily confirmed cases reported in Malaysia using training data of observed cases from Jan 22 to Mar 31, 2020. Subsequently, they validated using data on cases from Apr 1 to Apr 17, 2020, deducing that the ARIMA(0,1,0) model produced the best fit to the observed data. Ding et al. (2020) analyzed the epidemic data from Feb 24 to Mar 30, 2020, in Italy, based on ARIMA model. They selected ARIMA(2,1,0) to fit the logarithmic sequence of cumulative diagnoses. Hernandez-Matamoros et al. (2020) constructed a model for 145 countries, which are distributed in 6 geographic regions using the ARIMA parameters, the population per 1M people, the number of cases, and polynomial functions, the study period was until Apr 25. Dawoud (2020) Applied classical ARIMA together with the k^th^ MA - ARIMA to model the COVID-19 cumulative confirmed cases in Palestine from Mar 5, 2020, through Aug 27, 2020. He inferred that ARIMA (1,2,4) and 5th EWMA-ARIMA (2,2,3) were the best models. Duong et al. (2020) applied the ARIMA model for the total daily confirmed cases worldwide from Jan 21, 2020, to Mar 16, 2020. They found that ARIMA(1,2,1) could describe and predict the epidemiological trend of COVID-19. Roy et al.(2020) used ARIMA to fit the COVID-2019 disease from Jan 26, 2020, to May 9, 2020, in Indian. They selected ARIMA(1,0,2) model to fit the sequence of diagnoses. Verma et al. (2020) developed some models based on ARIMA and FUZZY time series methodology to forecast the COVID-19 infection, mortality, and recovery in India throughout the phase between Mar 2, 2020, to May 17, 2020. They deduced that the ARIMA and FUZZY time series models’ forecasts would be useful for the decision-makers.

The main objective of this article is to model confirmed, recovery, and death cases of COVID-19 using classical ARIMA besides the three types of k^th^ Moving Average-ARIMA (k^th^ MA-ARIMA), including k^th^ Simple Moving Average-ARIMA (k^th^ SMA-ARIMA), k^th^ Weighted Moving Average-ARIMA (k^th^ WMA-ARIMA) and k^th^ Exponential Weighted Moving Average-ARIMA (k^th^ EWMA-ARIMA) in the GCC countries. This study starts from the first case of coronavirus reported in each GCC country to Nov 30, 2020. This article’s main contribution is that it considers the only study that used the classical ARIMA together with k^th^ SMA-ARIMA, k^th^ WMA-ARIMA, and k^th^ EWMA-ARIMA to model the three variables confirmed, recovery, and death cases of COVID-19 in the GCC countries.

The organization of the paper is as follows. Section 2 describes the study area and data collection. Section 3 briefs the methodology used in the study. The article ends with the results and discussion in Section 4, and conclusions in Section 5.

## 2. Study Area and Data collection

To achieve this study’s objectives, all six countries within the GCC were included (Saudi Arabia, United Arab Emirates, Qatar, Kuwait, Bahrain, and Oman). The sample data consist of daily reported COVID-19 cases of 3 variables involving confirmed, recovery, and deaths in each country. The data cover the period starting from the first confirmed case of COVID-19 reported in each country to Nov 30, 2020. The data extracted from the WHO situation reports, Sehhty website, and Wikipedia.

## 3. Methodology

This paper’s main goal is to model 3 variables involving daily confirmed, recovery, and death cases in GCC countries using classical ARIMA besides the three types of k^th^ MA-ARIMA including k^th^ SMA-ARIMA, k^th^ WMA-ARIMA, and k^th^ EWMA-ARIMA. Therefore, this section investigates each of these models, discussing model building and model evaluation.

### 3.1. ARIMA Model

ARIMA model, which was developed by Box and Jenkins (1994), is a statistical model that uses time series data to study the trend and generate future forecasting of time series data.

For a given non-stationary time series *X*_*t*_, the classical *ARIMA* (*p, d, q*) model is defined as

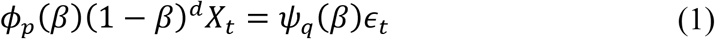

Where *β* is the backward shift operator, (1 − *β*)^*d*^ is the difference filter, *d* is a number of times, need to differentiate *X*_*t*_ to make the data stationary, *p* is the order of autoregression, *q* is the order of moving average, *ϕ*_*p*_(*β*) = 1 − *ϕ*_1_*β* − *ϕ*_2_*β*^2^ − … − *ϕ*_*p*_*β*^*p*^, *ψ*_*q*_(*β*) = 1 + *ψ*_1_*β* + *ψ*_2_*β*^2^ + ⋯ + +*ψ*_*q*_*β*^*q*^ and *ϵ*_*t*_ *∼ N*(0,1).

ARIMA model is a generalized model that integrates the autoregressive model *AR*(*p*) and the moving average model *MA*(*q*), ARIMA models that do not require differencing are considered as ARMA models, therefore model (1) can be expressed as polynomials of autoregressive *AR*(*p*), residuals *MA*(*q*), and a combination of them *ARMA*(*p, q*) as

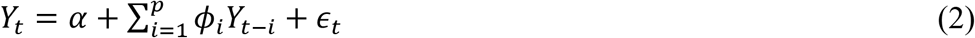

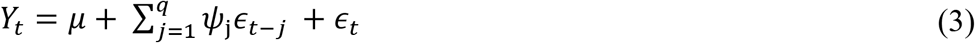

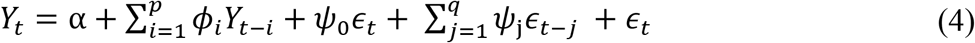

### 3.2. The k^th^ Moving Average-ARIMA Time Series Models

k^th^ Moving average ARIMA model technique (k^th^ MA-ARIMA) proposed by Shih and Tsokos (2008) and Tsokos (2010). This model is based on modifying a given time series into a new k-time moving average time series, and then developing the autoregressive integrated moving average (ARIMA) model using the Box and Jenkins method. Once the new time series’s forecasting model is built, a back-shift operator is applied to obtain estimates of the original phenomenon. The k^th^ MA-ARIMA involves k^th^ SMA-ARIMA, k^th^ WMA-ARIMA, and k^th^ EWMA-ARIMA.

#### 3.2.1. The k^th^ SMA-ARIMA Model

The k^th^ SMA-ARIMA process of a time series *x*_*t*_ and it is the corresponding back-shift operator are defined, respectively, by

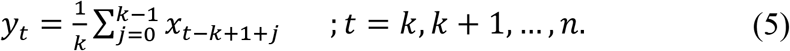

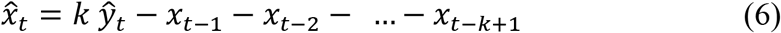

#### 3.2.2. The k^th^ WMA-ARIMA Model

The k^th^ WMA-ARIMA process of a time series *x*_*t*_ and it is the corresponding back-shift operator are given, respectively, as

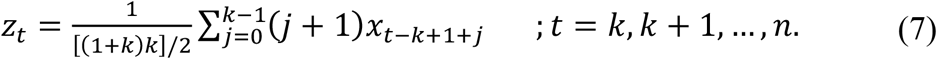

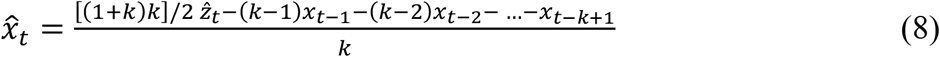

#### 3.2.3. The k^th^ EWMA-ARIMA Model

The k^th^ EWMA-ARIMA process of a time series *x*_*t*_ and it is the corresponding back-shift operator are computed, respectively, as

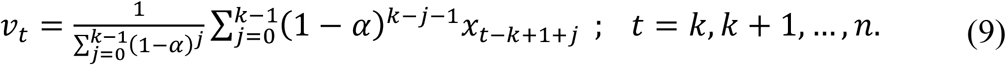

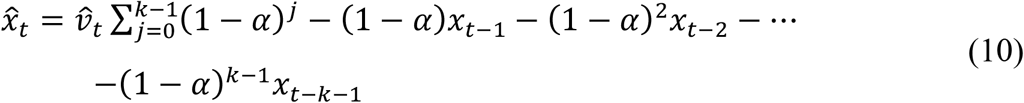

### 3.3. Model selection criteria

Model selection criteria are rules used to select a [statistical model](https://www.statlect.com/glossary/statistical-model) among a set of candidate models based on the observed data. The Akaike information criterion (AIC) is a widely used model selection tool due to its computational simplicity and effective performance in many modeling frameworks. The AIC is given as (Akaike, 1974)

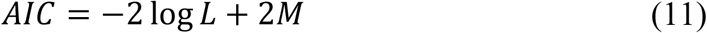

Where *L* is the likelihood of the model and *M* is the total number of estimated parameters in the model. A good model is the one that has the minimum AIC among all other models.

### 3.4. Measures of forecast accuracy

The most popular measure of forecast accuracy in univariate time series data is the Root Mean Square Error (RMSE) proposed by Hyndman and Koehler (2006). The RMSE is computed as

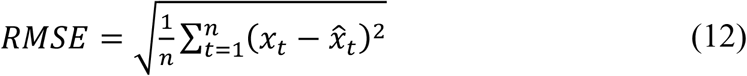

where *x*_*t*_ and 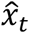 are the actual and predicted values at time *t*, respectively, and *n* is a sequence of time points. The lower value of RMSE indicates better calibration and, therefore, better performance.

### 3.5. Checking model’s goodness of fit

After the ARIMA or k^th^ MA-ARIMA model, which is considered appropriate among the alternatives, is put in place, it can be tested for a goodness fit, which entails testing its efficiency. The model is assumed to be a good fit if the residuals are approximately equal to the white noise. The essential tools are the plots of ACF and PACF. The Box-Ljung test is a diagnostic tool used to test the lack of fit of a time series model. This test is applied to the residuals of a time series after fitting an ARIMA or k^th^ MA-ARIMA model to the data. The test examines *m* autocorrelations of the residuals. The null and alternative hypothesis for this test is

*H*_0_: The model does not exhibit a lack of fit, or there is no serial correlation among *m* lags

*H*_1_: The model exhibits a lack of fit, or the residuals are approximately equal to the white noise.

## 4. Results and Discussion

This section first demonstrates summary statistics for the three variables, confirmed, recovery, and death cases in each GCC country, then reports and discusses the results obtained from applying the ARIMA and k^th^ MA-ARIMA models on these variables.

### 4.1. Summary Statistics for COVID-19 confirmed, recovery and death cases

Table 1 shows the summary statistics measures, including mean and standard deviation of the confirmed, recovery, and death cases of COVID-19 among the GCC countries. Moreover, Table 1 also demonstrates the prevalence of confirmed cases per 100000 population for the first four weeks.

**Table 1.**
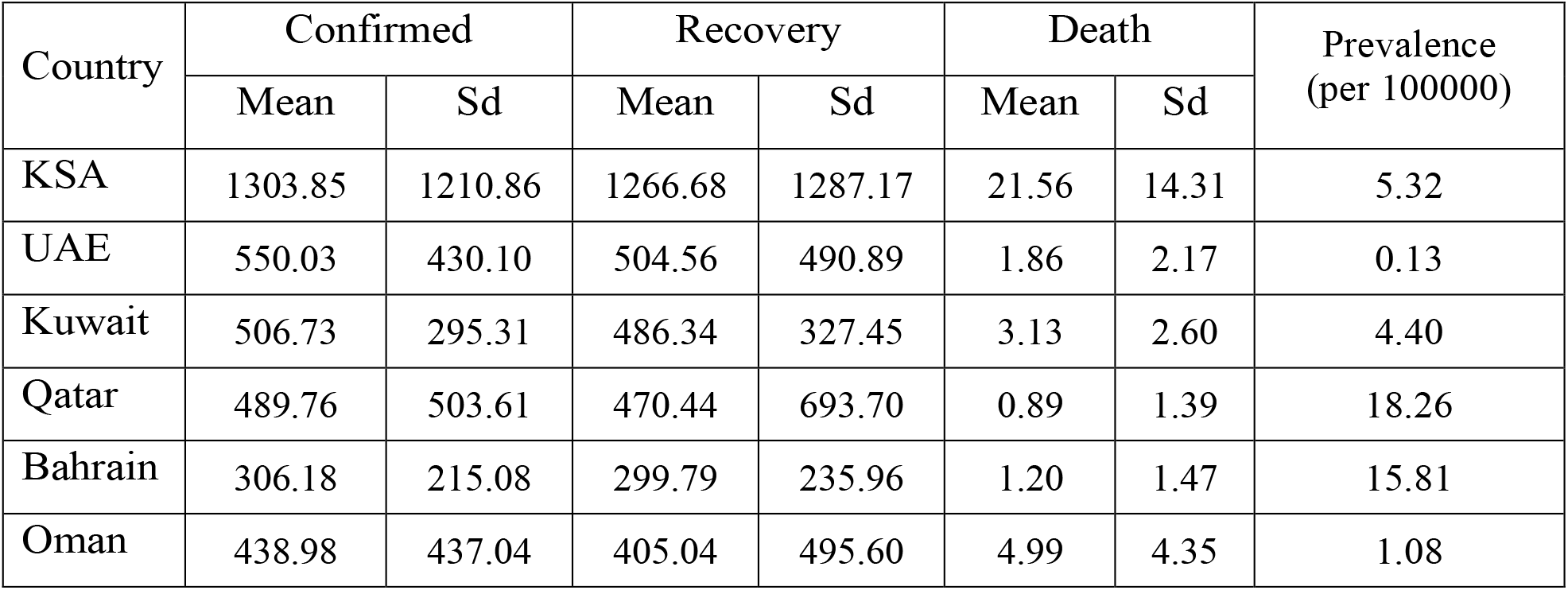
Mean and standard deviation of confirmed, recovery and death cases in GCC countries among the study periods and prevalence of cumulative confirmed cases.

| Country | Confirmed |  | Recovery |  | Death |  | Prevalence<br>(per 100000) |
| --- | --- | --- | --- | --- | --- | --- | --- |
|  | Mean | Sd | Mean | Sd | Mean | Sd |  |
| KSA | 1303.85 | 1210.86 | 1266.68 | 1287.17 | 21.56 | 14.31 | 5.32 |
| UAE | 550.03 | 430.10 | 504.56 | 490.89 | 1.86 | 2.17 | 0.13 |
| Kuwait | 506.73 | 295.31 | 486.34 | 327.45 | 3.13 | 2.60 | 4.40 |
| Qatar | 489.76 | 503.61 | 470.44 | 693.70 | 0.89 | 1.39 | 18.26 |
| Bahrain | 306.18 | 215.08 | 299.79 | 235.96 | 1.20 | 1.47 | 15.81 |
| Oman | 438.98 | 437.04 | 405.04 | 495.60 | 4.99 | 4.35 | 1.08 |

Based on Table 1, it is observed that KSA has the highest mean of confirmed cases (1303.85) with a standard deviation of (1210.86), followed by UAE, Kuwait, and Oman; on the other side, Bahrain has the lowest mean (306.18) with a standard deviation of (215.08). For recovery cases, KSA has the highest mean, followed by UAE, Kuwait, Qatar, and Oman, but Bahrain has the lowest one. KSA has the highest mean of reported death cases, followed by Oman, Kuwait. On the other hand, Qatar has the lowest one. It can be also seen that in the first 4 weeks of COVID-19 outbreak, Qatar and Bahrain have the highest prevalence of confirmed cases of 18 and 16 infected persons per 1000000, respectively. In contrast, UAE and Oman have the lowest ones of 1 and 1.1 per 1000000, respectively (see Figure 1).

**Figure 1.**
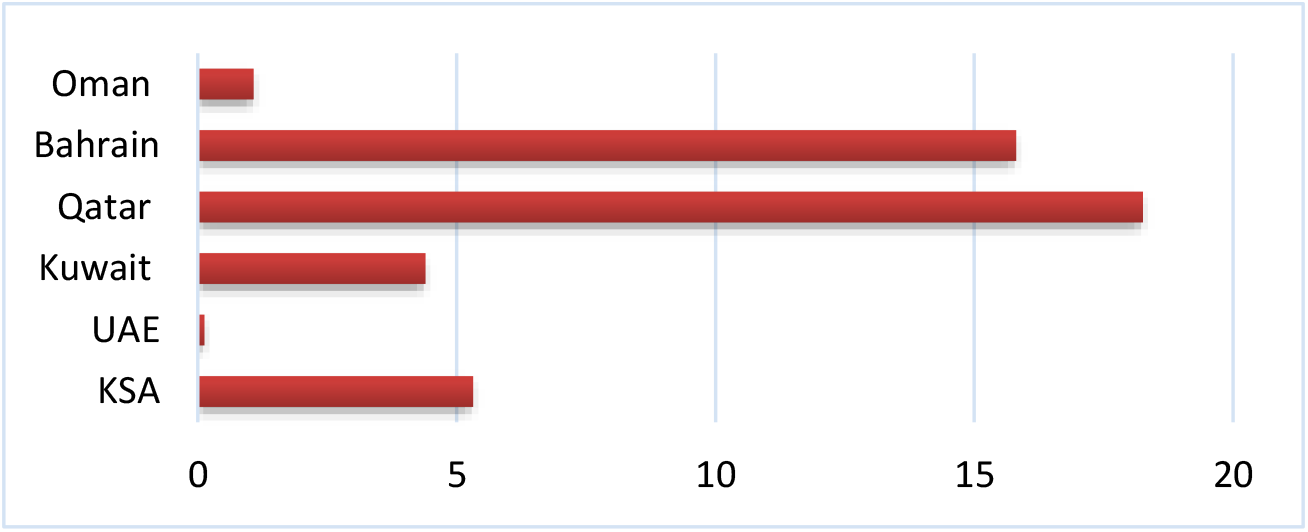
The prevalence of confirmed cases per 100000 population for the first four weeks in GCC countries

### 4.2. Prediction model for COVID-19 confirmed, recovery and death cases

This paper uses the time series, daily COVID-19 confirmed, recovery, and death cases in each GCC country. Therefore, we have a time series presented as follows:

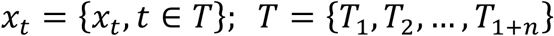

where *x*_*t*_ represents the confirmed, recovery or death cases at day *t* and *T*_1_ denotes the date of the first case of COVID-19 detected in a given country. The time-series plot of the daily COVID-19 confirmed, recovery, and death cases for GCC countries is presented in Figure 2, Figure 3, and Figure 4, respectively.

**Figure 2.**
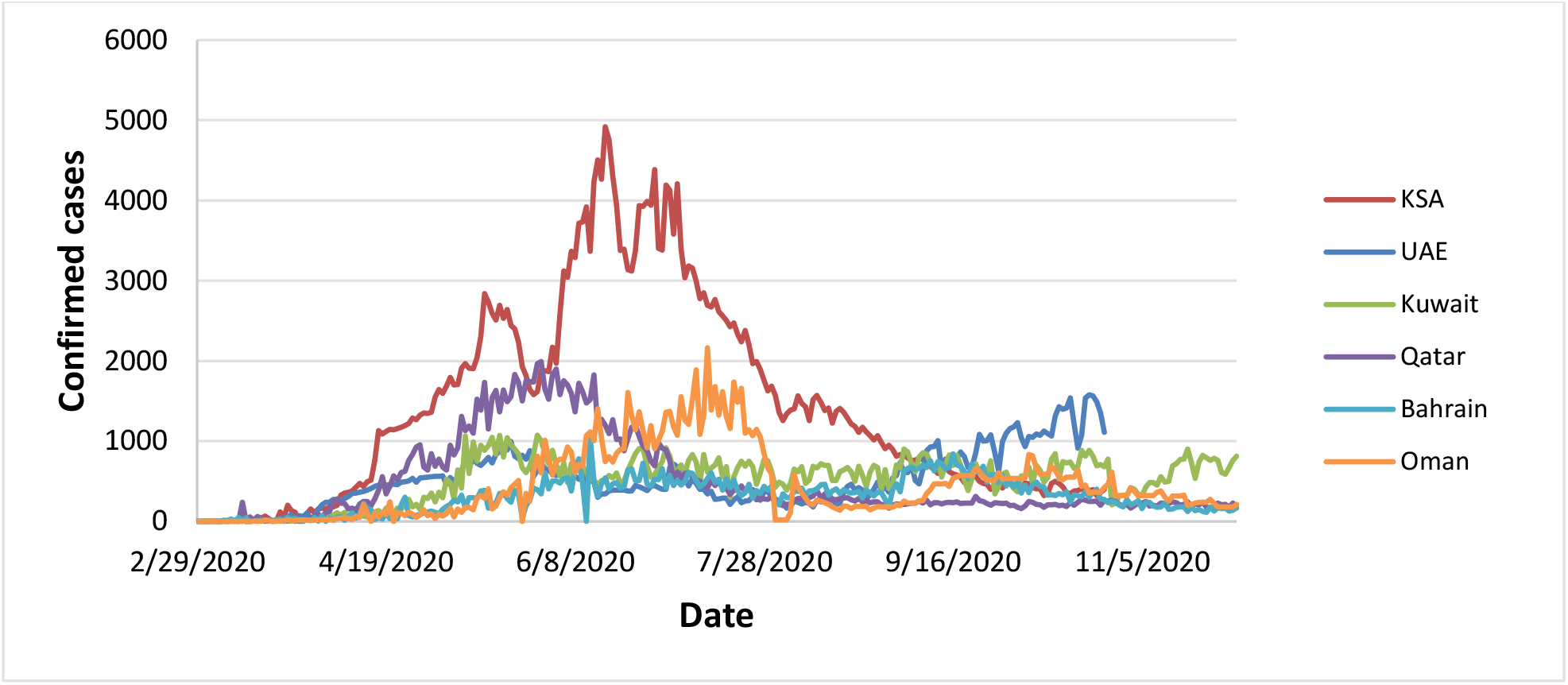
Time-series of daily COVID-19 confirmed cases in each country

**Figure 3.**
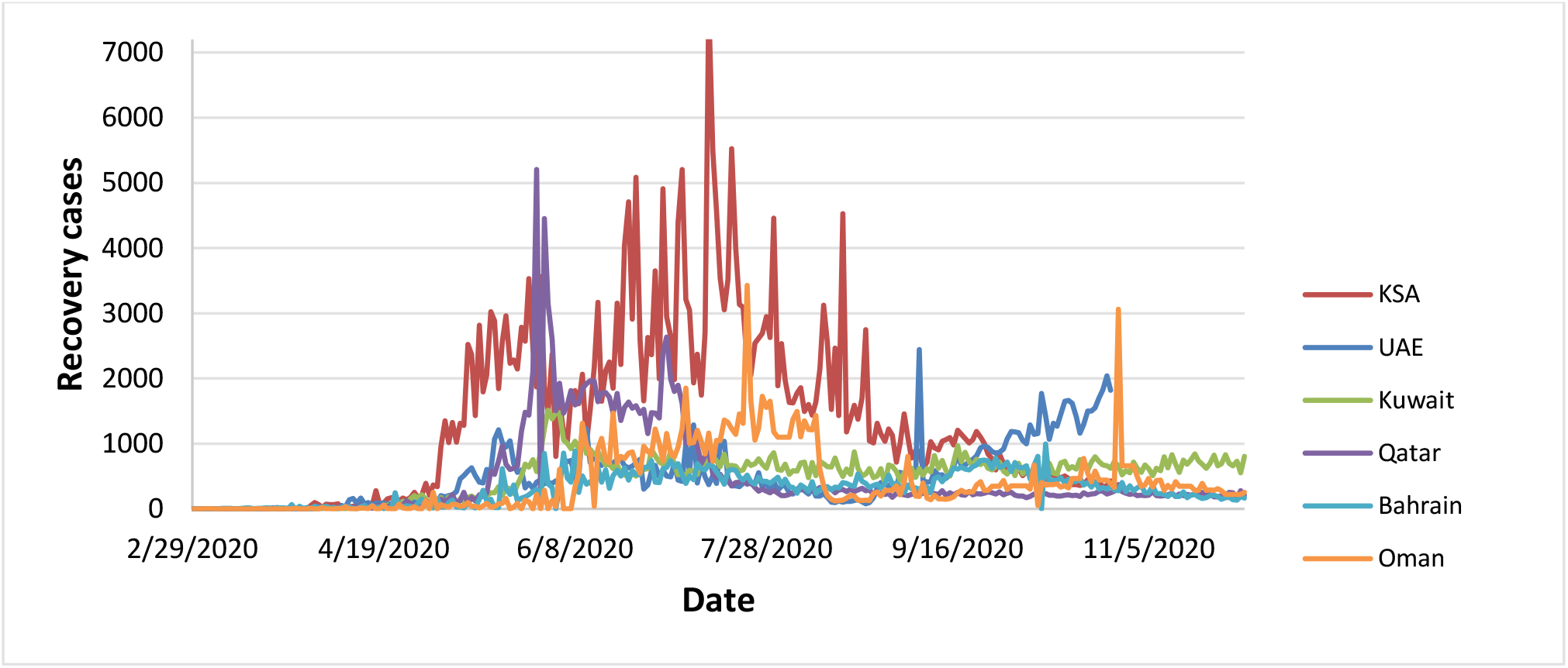
Time-series of daily COVID-19 recovery cases in each country

**Figure 4.**
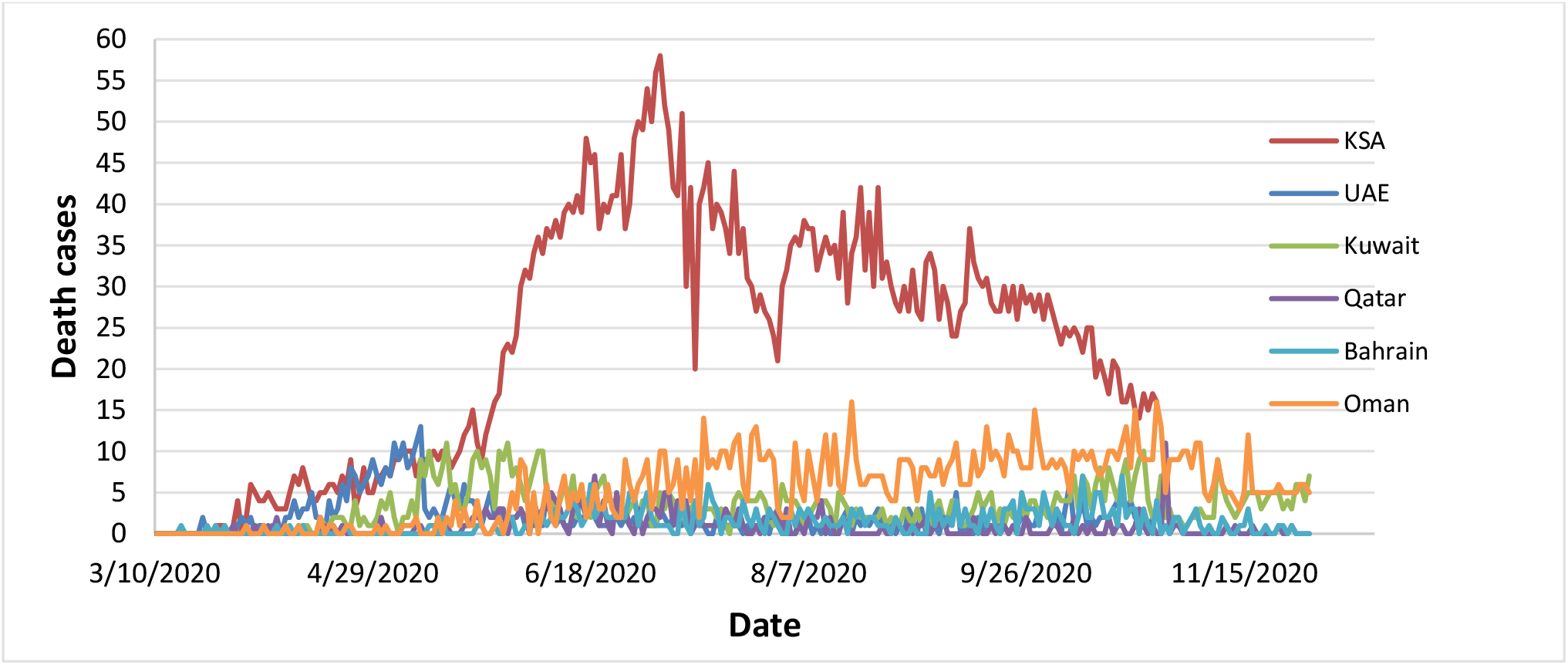
Time-series of daily COVID-19 death cases in each country

### 4.3. Prediction Method

To compute the best parameters estimates of ARIMA, k^th^ SMA-ARIMA, k^th^ WMA-ARIMA, and k^th^ EWMA-ARIMA models, these models were fitted for 90% of the available data in each country which is called the in-sample forecast or training data and the remaining 10% was used for the out-of-sample forecast or testing the model. The AIC of Eq. (11) was applied to the training data as a criterion method to select the best model. Furthermore, the statistical measure RMSE of Eq.(12) was utilized for testing data, and the model with the minimum AIC and minimum RMSE was selected. The calculations were performed using R studio version 1.2.5033 and EViews 10.

### 4.4. ARIMA model for COVID-19 confirmed, recovery and death cases

To check whether the daily COVID-19 confirmed, recovery and death cases time series in each country were stationary; we carried on ADF root test. The results of the ADF unit root test are demonstrated in Table A.1 in the Appendix. Based on Table A.1, we conclude that all variables are stationary with constant and trend at first differences throughout the study period; therefore, the ARIMA model can be done. After the stationarity of the confirmed, recovery, and death cases time series in each country were determined, the best ARIMA model that fit these 3 variables well for training data with the minimum AIC and lowest RMSE were selected. Table 2 summarizes the best ARIMA model for the confirmed recovery and death cases in each country and their corresponding RMSE and AIC.

**Table 2:** The best ARIMA model fitting confirmed, recovery and death cases and their corresponding RMSE, and AIC.

| Country | Confirmed cases |  |  | Recovery cases |  |  | Death cases |  |  |
| --- | --- | --- | --- | --- | --- | --- | --- | --- | --- |
|  | ARIMA<br>(p,d,q) | RMSE | AIC | ARIMA<br>(p,d,q) | RMSE | AIC | ARIMA<br>(p,d,q) | RMSE | AIC |
| KSA | (2,1,3) | 52.226 | 3607.47 | (1,1,1) | 37.38 | 4326.68 | (1,1,2) | 2.175 | 1448.66 |
| UAE | (2,1,2) | 90.93 | 3602.49 | (0,1,1) | 171.43 | 4117.92 | (0,1,1) | 1.774 | 1064.88 |
| Kuwait | (3,1,2) | 98.51 | 3480.61 | (0,1,1) | 105.97 | 3401.79 | (2,1,3) | 1.715 | 1087.25 |
| Qatar | (2,1,3) | 26.09 | 3353.06 | (1,1,2) | 40.39 | 3991.72 | (0,1,1) | 0.566 | 888.19 |
| Bahrain | (3,1,0) | 30.50 | 3354.88 | (1,1,2) | 41.39 | 3474.46 | (2,1,3) | 0.913 | 908.01 |
| Oman | (2,1,2) | 48.52 | 3610.16 | (0,1,1) | 97.54 | 3961.79 | (0,1,1) | 2.313 | 1257.95 |

Based on the results in Table 2, we observed that ARIMA (2,1,3), ARIMA (1,1,1), and ARIMA (1,1,2) consider as the best models to fit the confirmed, recovery, and death cases of COVID-19 in Saudi Arabia, respectively; these models have the minimum AICs (3607.47, 4326.68, 1448.66) and lowest RMSEs (52.226, 37.38, 2.175) values among all models. These results imply that ARIMA (2,1,3), ARIMA (1,1,1), and ARIMA (1,1,2) are more efficient than other ARIMA models with *p* + *q ≤* 5 for in-sample forecasts. Consequently, a new confirmed, recovery, and death case can be interpreted based on the current case and the most recent change of the COVID-19 trend. The remaining results of Table 2 can be interpreted in the same manner.

### 4.5. k^th^ MA-ARIMA model for COVID-19 confirmed, recovery and death cases

We can summarize the process of developing the k^th^ SMA-ARIMA, k^th^ WMA-ARIMA, and k^th^ EWMA-ARIMA models as follows:

1. Transforming the original time series *x*_*t*_ into the new one (*y*_*t*_, *z*_*t*_, *v*_*t*_) for *k* = 2,3, …, 5 by using Eq. (5), Eq. (7), and Eq. (9), respectively.
2. Checking the stationary of time series (*y*_*t*_, *z*_*t*_, *v*_*t*_) using the ACF test until we achieve stationarity.
3. Applying the classical *ARIMA*(*p, d, q*) for the *y*_*t*_, *z*_*t*_ or *v*_*t*_ determined in step 2, where *p* + *q ≤* 5.
4. Compute the AIC for each model, and choose the one with the smallest AIC.
5. Solve the estimates of the original time series (fitted values) by using the back-shift operator of Eq. (6), Eq. (8), and Eq. (10), respectively.
6. Computing the RMSE for each model, and choose one with the smallest RMSE.

After taking the first differences of the transformed data to make it stationarity, we fitted 72 models for each type of the 3 k^th^ MA-ARIMA models (6 countries *×* 3 variables *×* 4 values of *k* (*k* = 2,3,4, 5)). The best 18 out of 72 different combinations of k^th^ SMA-ARIMA, k^th^ WMA-ARIMA, and k^th^ EWMA-ARIMA models fitting the confirmed, recovery and death cases of COVID-19 well with the corresponding RMSE and AIC for each country are presented in Table 3, Table 4, and Table 5, respectively.

**Table 3:** The best k^th^ SMA-ARIMA models fitting confirmed, recovery and death cases and their corresponding RMSE and AIC in each country.

| Country | Confirmed cases |  |  | Recovery cases |  |  | Death cases |  |  |
| --- | --- | --- | --- | --- | --- | --- | --- | --- | --- |
| | $k^{\text{th}}$<br>SMA-<br>ARIMA<br>(p,d,q) | RMSE | AIC | $k^{\text{th}}$<br>SMA-<br>ARIMA<br>(p,d,q) | RMSE | AIC | $k^{\text{th}}$<br>SMA-<br>ARIMA<br>(p,d,q) | RMSE | AIC |
| KSA | 2-(2,1,3) | 50.96 | 3226.27 | 2-(2,1,2) | 41.96 | 3942.94 | 2-(3,1,1) | 2.25 | 1073.40 |
| UAE | 4-(2,1,3) | 88.78 | 2772.99 | 5-(1,1,4) | 156.26 | 3142.70 | 3-(0,1,3) | 1.741 | 402.94 |
| Kuwait | 4-(0,1,4) | 95.86 | 2701.30 | 2-(0,1,2) | 99.72 | 3009.86 | 3-(1,1,3) | 1.570 | 485.63 |
| Qatar | 5-(1,1,4) | 26.94 | 2470.52 | 3-(3,1,1) | 39.23 | 3427.90 | 2-(0,1,2) | 0.623 | 509.76 |
| Bahrain | 4-(2,1,3) | 29.54 | 2574.88 | 3-(1,1,4) | 41.36 | 2845.80 | 3-(0,1,3) | 0.931 | 298.96 |
| Oman | 4-(0,1,4) | 40.77 | 2824.50 | 5-(1,1,4) | 112.04 | 3083.97 | 4-(0,1,4) | 2.321 | 496.58 |

**Table 4:**
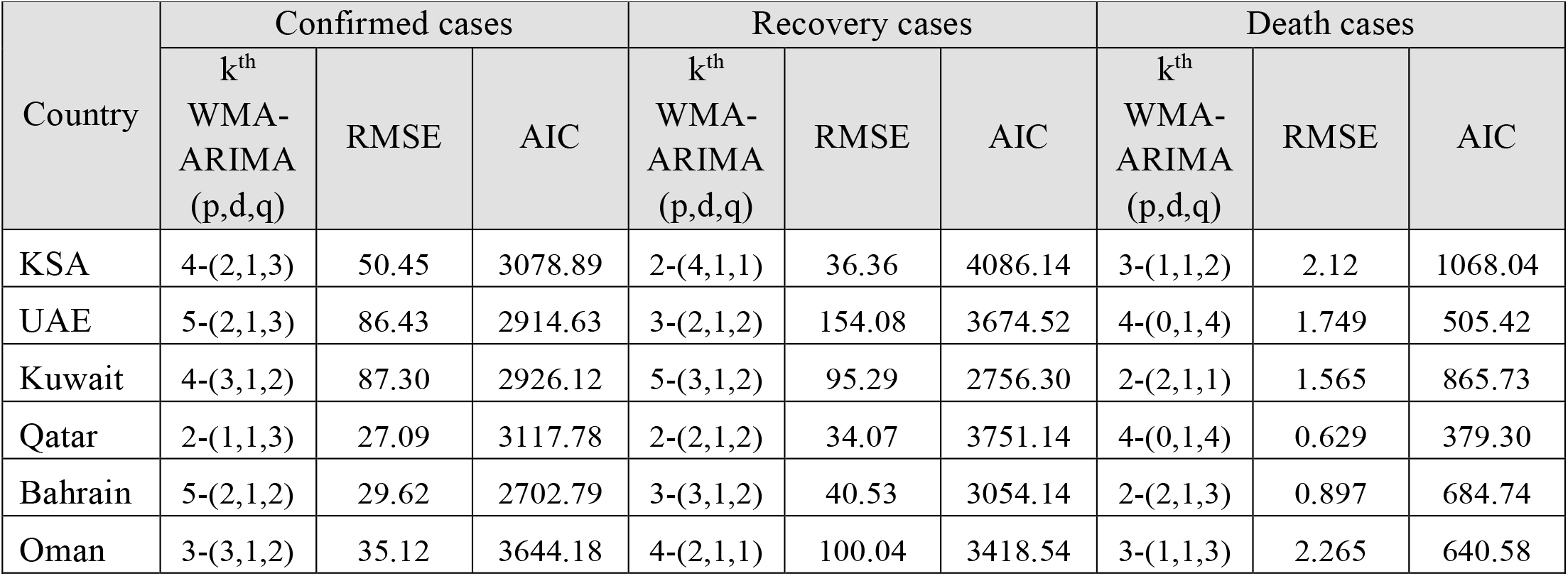
The best k^th^ WMA-ARIMA models fitting confirmed, recovery and death cases and their corresponding RMSE and AIC in each country.

**Table 5:** The best k^th^ EWMA-ARIMA models fitting confirmed, recovery and death cases and their corresponding RMSE and AIC in each country.

| Country | Confirmed cases |  |  | Recovery cases |  |  | Death cases |  |  |
| --- | --- | --- | --- | --- | --- | --- | --- | --- | --- |
| | $k^{\text{th}}$<br>EWMA-<br>ARIMA<br>(p,d,q) | RMSE | AIC | $k^{\text{th}}$<br>EWMA-<br>ARIMA<br>(p,d,q) | RMSE | AIC | $k^{\text{th}}$<br>EWMA-<br>ARIMA<br>(p,d,q) | RMSE | AIC |
| KSA | 4-(2,1,3) | 50.24 | 3153.81 | 2-(3,1,2) | 37.03 | 4158.79 | 3-(1,1,2) | 2.120 | 1138.92 |
| UAE | 5-(2,1,3) | 87.39 | 2995.63 | 5-(2,1,3) | 172.24 | 3495.32 | 4-(0,1,4) | 1.750 | 589.17 |
| Kuwait | 2-(2,1,2) | 97.02 | 3314.08 | 3-(2,1,2) | 95.34 | 3069.93 | 3-(3,1,1) | 1.599 | 775.04 |
| Qatar | 2-(1,1,3) | 27.03 | 3184.76 | 5-(1,1,4) | 33.79 | 3413.73 | 3-(1,1,1) | 0.578 | 575.81 |
| Bahrain | 2-(3,1,0) | 29.16 | 3182.10 | 3-(3,1,2) | 41.08 | 3128.73 | 5-(1,1,1) | 0.911 | 377.85 |
| Oman | 3-(2,1,3) | 34.71 | 3265.76 | 4-(1,1,1) | 94.76 | 3493.31 | 3-(0,1,3) | 2.253 | 938.40 |

Depending on the results in Table 3, it can be concluded that the 2^nd^ SMA-ARIMA(2,1,3), 2^nd^ SMA-ARIMA(2,1,2), and 2^nd^ SMA-ARIMA(3,1,1) were selected as the best models to fit the confirmed, recovery, and death cases of COVID-19 in Saudi Arabia, respectively. These models have the minimum AICs (3223.7, 3942.94, 1072.40) and lowest RMSEs (50.96, 41.96, 2.25) values among all SMA-ARIMA models. These results imply that the selected models are more efficient than other SMA-ARIMA models with *k* = 2,3,4, 5 and *p* + *q ≤* 5 for in-sample forecasts. Accordingly, a new confirmed, recovery, and death case can be interpreted based on the current case and the most recent change of the COVID-19 trend. The remaining results of Table 3 and the outputs in Table 4 and Table 5 can be interpreted in the same manner.

Table 6 reviews the best models among the k^th^ MA-ARIMA models based on the smallest RMSE. In contrast, Table 7 shows the best models among classical ARIMA besides the k^th^ MA-ARIMA based on the smallest RMSE.

**Table 6:** The best k^th^ MA-ARIMA models fit confirmed, recovery, and death cases in each country.

| Country | Confirmed cases |  | Recovery cases |  | Death cases |  |
| --- | --- | --- | --- | --- | --- | --- |
|  | Model | RMSE | Model | RMSE | Model | RMSE |
| KSA | 4<sup>th</sup> EWMA-ARIMA<br>(2,1,3) | 50.24 | 2<sup>nd</sup> WMA-ARIMA<br>(4,1,1) | 36.36 | 3<sup>rd</sup> WMA-ARIMA<br>(1,1,2) | 2.120 |
| UAE | 5<sup>th</sup> WMA-ARIMA<br>(2,1,3) | 86.43 | 3<sup>rd</sup> WMA-ARIMA<br>(2,1,2) | 154.08 | 3<sup>rd</sup> SMA-ARIMA<br>(0,1,3) | 1.741 |
| Kuwait | 4<sup>th</sup> WMA-ARIMA<br>(3,1,2) | 87.30 | 5<sup>th</sup> WMA-ARIMA<br>(3,1,2) | 95.29 | 2<sup>nd</sup> WMA-ARIMA<br>(2,1,1) | 1.565 |
| Qatar | 5<sup>th</sup> SMA-ARIMA<br>(1,1,4) | 26.94 | 5<sup>th</sup> EWMA-ARIMA<br>(1,1,4) | 33.79 | 3<sup>rd</sup> EWMA-ARIMA<br>(1,1,1) | 0.578 |
| Bahrain | 2<sup>nd</sup> EWMA-<br>ARIMA (3,1,0) | 29.16 | 3<sup>rd</sup> EWMA-ARIMA<br>(3,1,2) | 41.08 | 2<sup>nd</sup> WMA-ARIMA<br>(2,1,3) | 0.897 |
| Oman | 3<sup>rd</sup> EWMA-ARIMA<br>(2,1,3) | 34.71 | 4<sup>th</sup> EWMA-ARIMA<br>(1,1,1) | 94.76 | 3<sup>rd</sup> EWMA-ARIMA<br>(0,1,3) | 2.253 |

**Table 7:** The best models among the classical ARIMA and k^th^ MA-ARIMA fitting confirmed, recovery, and death cases in each country.

| Country | Confirmed cases |  | Recovery cases |  | Death cases |  |
| --- | --- | --- | --- | --- | --- | --- |
|  | Model | RMSE | Model | RMSE | Model | RMSE |
| KSA | 4<sup>th</sup> EWMA-ARIMA (2,1,3) | 50.24 | 2<sup>nd</sup> WMA-ARIMA (4,1,1) | 36.36 | 3<sup>rd</sup> WMA-ARIMA (1,1,2) | 2.120 |
| UAE | 5<sup>th</sup> WMA-ARIMA (2,1,3) | 86.43 | 3<sup>rd</sup> WMA-ARIMA (2,1,2) | 154.08 | 3<sup>rd</sup> SMA-ARIMA (0,1,3) | 1.741 |
| Kuwait | 4<sup>th</sup> WMA-ARIMA (3,1,2) | 87.30 | 5<sup>th</sup> WMA-ARIMA (3,1,2) | 95.29 | 2<sup>nd</sup> WMA-ARIMA (2,1,1) | 1.565 |
| Qatar | ARIMA(2,1,3) | 26.09 | 5<sup>th</sup> EWMA-ARIMA (1,1,4) | 33.79 | ARIMA(0,1,1) | 0.566 |
| Bahrain | 2<sup>nd</sup> EWMA-ARIMA (3,1,0) | 29.16 | 3<sup>rd</sup> EWMA-ARIMA (3,1,2) | 41.08 | 2<sup>nd</sup> WMA-ARIMA (2,1,3) | 0.897 |
| Oman | 3<sup>rd</sup> EWMA-ARIMA (2,1,3) | 34.71 | 4<sup>th</sup> EWMA-ARIMA (1,1,1) | 94.76 | 3<sup>rd</sup> EWMA-ARIMA (0,1,3) | 2.253 |

After identifying the best model within the classical ARIMA and k^th^ MA-ARIMA models fitting confirmed, recovery and death cases for each country (see: Table 7), the next step is to check the pattern followed by residuals from the specific model by plotting the ACF of the residuals and conducting the Box-Ljung test to examine the goodness of fit for each models. Figures (5.a.1 to 5.c6) show ACF plots for all the best models located in Table 7, while Table 8 demonstrates the outputs of the Box-Ljung test.

**Table 8:** The Box-Ljung test applied for the best models among ARIMA and k^th^ MA -ARIMA models fitting confirmed, recovery and death cases in each country.

| Country | Confirmed cases |  |  | Recovered cases |  |  | Death cases |  |  |
| --- | --- | --- | --- | --- | --- | --- | --- | --- | --- |
|  | Model | Q* | P-value | Model | Q* | P-value | Model | Q* | P-value |
| KSA | 4<sup>th</sup> EWMA-ARIMA (2,1,3) | 34.135 | 0.276 | 2<sup>nd</sup> WMA-ARIMA (4,1,1) | 35.38 | 0.229 | 3<sup>rd</sup> WMA-ARIMA (1,1,2) | 39.83 | 0.108 |
| UAE | 5<sup>th</sup> WMA-ARIMA (2,1,3) | 140.12 | <0.001 | 3<sup>rd</sup> WMA-ARIMA (2,1,2) | 17.03 | 0.972 | 3<sup>rd</sup> SMA-ARIMA (0,1,3) | 33.28 | 0.311 |
| Kuwait | 4<sup>th</sup> WMA-ARIMA (3,1,2) | 33.71 | 0.293 | 5<sup>th</sup> WMA-ARIMA (3,1,2) | 26.22 | 0.664 | 2<sup>nd</sup> WMA-ARIMA (2,1,1) | 40.57 | 0.094 |
| Qatar | ARIMA (2,1,3) | 66.83 | 0.0001 | 5<sup>th</sup> EWMA-ARIMA (1,1,4) | 27.91 | 0.575 | ARIMA (0,1,1) | 21.67 | 0.866 |
| Bahrain | 2<sup>nd</sup> EWMA-ARIMA (3,1,0) | 24.72 | 0.739 | 3<sup>rd</sup> EWMA-ARIMA (3,1,2) | 22.74 | 0.826 | 2<sup>nd</sup> WMA-ARIMA (2,1,3) | 33.80 | 0.289 |
| Oman | 3<sup>rd</sup> EWMA-ARIMA (2,1,3) | 34.37 | 0.266 | 4<sup>th</sup> EWMA-ARIMA (1,1,1) | 18.34 | 0.953 | 3<sup>rd</sup> EWMA-ARIMA (0,1,3) | 16.04 | 0.982 |

By looking at the ACF plots in all sub-Figures of Figure 5, it is observed that for the first 30 lags, most of the autocorrelations are inside the 95% confidence interval bounds indicating that they are white noise and normally distributed except ACF of Figure a2 and Figure a4 which have deviated a little from normality and randomized. The outputs of the Ljung-Box test in Table 8 confirm that there is no autocorrelation left on the residuals for all models in Table 7 except the two models concerning confirmed cases in UAE and Qatar, and the null hypothesis that the residuals were white noise was not rejected and therefore, all models were exhibited goodness of fit. Thus, each model in Table 7 has passed the required checks and is ready for forecasting except the two models 5^th^ WMA-ARIMA(2,1,3) and ARIMA(2,1,3) corresponding to the confirmed cases in UAE and Qatar respectively.

**Figure 5.**
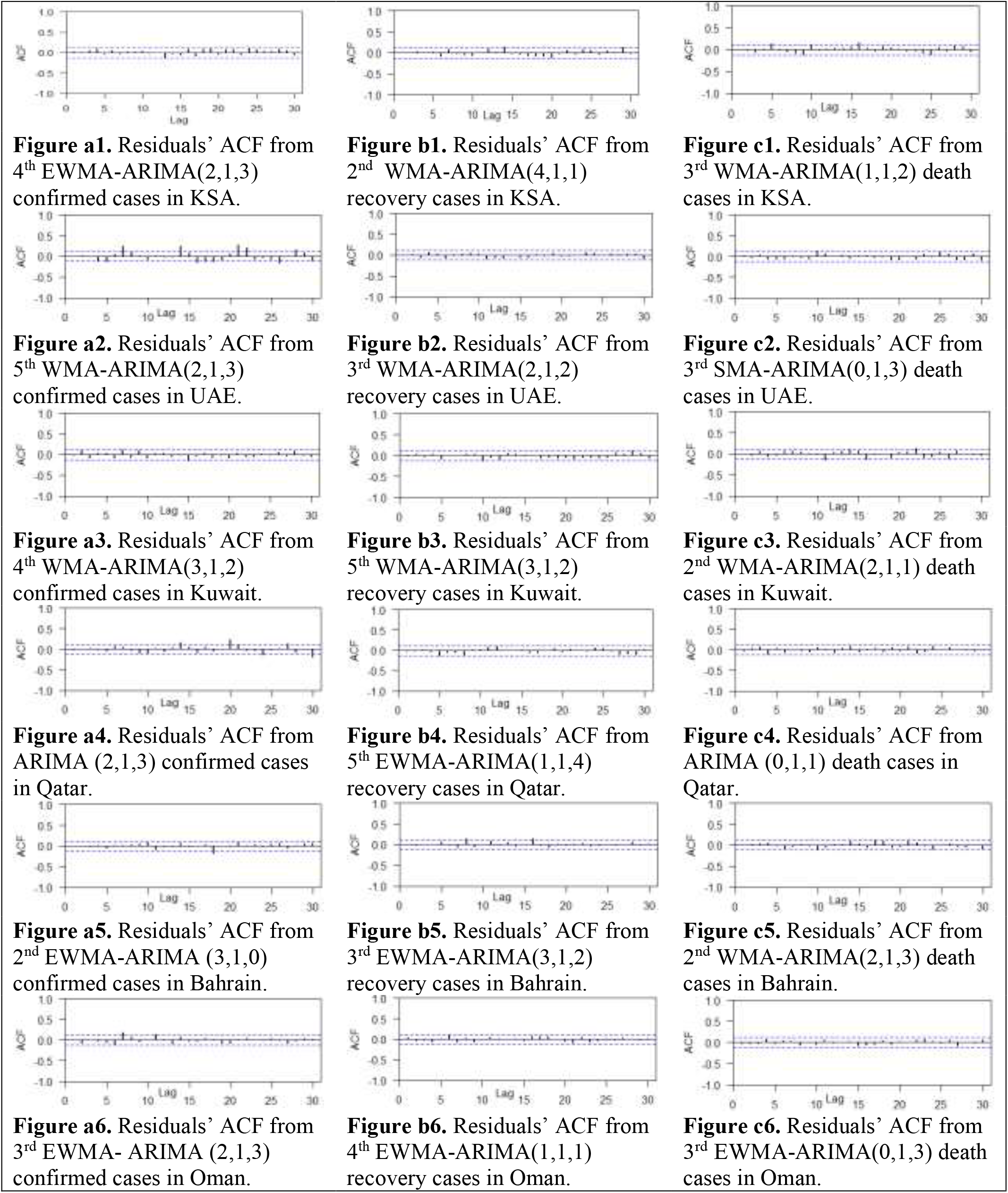
Residuals’ ACF from the best models confirmed, recovery and death cases in each country.

Tables 9, 10, and 11, respectively, demonstrate the forecasting result of confirmed, recovery and death cases for COVID-19 in each GCC country from Dec. 1, 2020 to Dec. 10, 2020 (10 values) based on each corresponding model listed in Table 7 (expected confirmed cases in UAE and Qatar). On the other hand, the suitable model for the confirmed cases in UAE and Qatar is the cubic linear regression model, and the estimated model for confirmed cases in UAE is

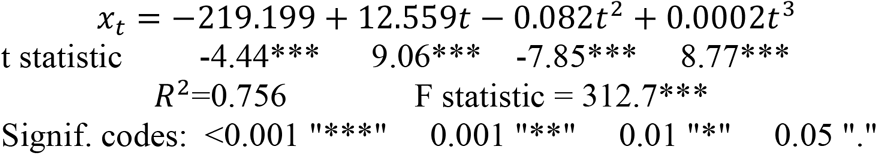

while the estimated cubic linear regression model for confirmed cases in Qatar is

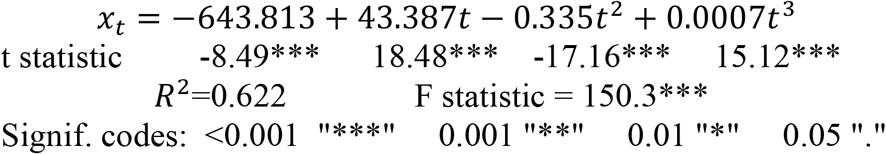

**Table 9.** Forecast values for daily COVID 19 confirmed cases from 01/12/2020 to 01/12/2020 based on the best models in each country.

| Date | KSA | UAE | Kuwait | Qatar | Bahrain | Oman |
| --- | --- | --- | --- | --- | --- | --- |
|  | 4<sup>th</sup> EWMA-ARIMA (2,1,3) | Cubic model | 4<sup>th</sup> WMA-ARIMA (3,1,2) | Cubic model | 2<sup>nd</sup> EWMA-ARIMA (3,1,0) | 3<sup>rd</sup> EWMA-ARIMA (2,1,3) |
| 01/12/2020 | 303.52 | 1583.99 | 803.50 | 479.74 | 155.91 | 191.46 |
| 02/12/2020 | 287.59 | 1601.79 | 685.79 | 498.82 | 144.36 | 173.99 |
| 03/12/2020 | 281.17 | 1619.79 | 655.00 | 518.40 | 153.60 | 177.02 |
| 04/12/2020 | 262.48 | 1637.98 | 675.22 | 538.49 | 154.80 | 192.57 |
| 05/12/2020 | 271.27 | 1656.38 | 733.28 | 559.08 | 151.29 | 201.04 |
| 06/12/2020 | 276.34 | 1674.98 | 764.02 | 580.18 | 151.06 | 199.70 |
| 07/12/2020 | 277.77 | 1693.78 | 771.26 | 601.79 | 152.75 | 189.79 |
| 08/12/2020 | 273.05 | 1712.79 | 738.17 | 623.94 | 152.55 | 179.45 |
| 09/12/2020 | 272.86 | 1732.01 | 691.25 | 646.60 | 151.84 | 177.85 |
| 10/12/2020 | 274.13 | 1751.43 | 666.33 | 669.79 | 152.04 | 185.72 |

**Table 10.** Forecast values for daily COVID 19 recovery cases from 01/12/2020 to 01/12/2020 based on the best models in each country.

| Date | KSA | UAE | Kuwait | Qatar | Bahrain | Oman |
| --- | --- | --- | --- | --- | --- | --- |
|  | 2<sup>nd</sup> WMA-ARIMA (4,11) | 5<sup>th</sup> SMA-ARIMA (1,1,4) | 5<sup>th</sup> WMA-ARIMA (3,1,2) | 5<sup>th</sup> EWMA-ARIMA (1,1,4) | 3<sup>rd</sup> EWMA-ARIMA (3,1,2) | 4<sup>th</sup> EWMA-ARIMA (1,1,1) |
| 01/12/2020 | 378.57 | 701.36 | 811.13 | 187.97 | 172.18 | 242.33 |
| 02/12/2020 | 412.09 | 745.99 | 746.88 | 204.85 | 161.61 | 243.34 |
| 03/12/2020 | 393.70 | 751.82 | 721.23 | 233.94 | 156.21 | 242.00 |
| 04/12/2020 | 401.12 | 748.71 | 719.37 | 217.73 | 155.42 | 242.09 |
| 05/12/2020 | 392.23 | 749.74 | 731.06 | 220.68 | 155.48 | 243.74 |
| 06/12/2020 | 392.91 | 741.44 | 750.24 | 210.21 | 158.32 | 244.05 |
| 07/12/2020 | 393.94 | 737.04 | 730.70 | 217.43 | 161.99 | 244.00 |
| 08/12/2020 | 395.69 | 738.03 | 729.42 | 217.93 | 164.28 | 244.11 |
| 09/12/2020 | 396.42 | 739.78 | 735.65 | 218.02 | 164.67 | 244.39 |
| 10/12/2020 | 396.53 | 742.16 | 735.75 | 216.93 | 163.28 | 244.48 |

**Table 11.** Forecast values for daily COVID 19 death cases from 01/12/2020 to 01/12/2020 based on the best models in each country.

| Date | KSA | UAE | Kuwait | Qatar | Bahrain | Oman |
| --- | --- | --- | --- | --- | --- | --- |
|  | 3<sup>rd</sup> WMA-ARIMA (1,1,2) | 3<sup>th</sup> SMA-ARIMA (0,1,3) | 3<sup>rd</sup> SMA-ARIMA (1,1,3) | ARIMA (0,1,1) | 2<sup>nd</sup> WMA-ARIMA (2,1,3) | 3<sup>rd</sup> WMA-ARIMA (0,1,3) |
| 01/12/2020 | 9.18 | 0.54 | 6.827 | 0.185 | 0.020 | 5.233 |
| 02/12/2020 | 12.23 | 3.18 | 4.734 | 0.185 | 0.476 | 5.419 |
| 03/12/2020 | 11.42 | 2.25 | 5.332 | 0.185 | 0.131 | 5.479 |
| 04/12/2020 | 11.06 | 0.54 | 4.984 | 0.185 | 0.362 | 5.402 |
| 05/12/2020 | 11.48 | 3.18 | 5.274 | 0.185 | 0.216 | 5.425 |
| 06/12/2020 | 11.40 | 2.25 | 5.222 | 0.185 | 0.306 | 5.433 |
| 07/12/2020 | 11.25 | 0.54 | 5.270 | 0.185 | 0.251 | 5.423 |
| 08/12/2020 | 11.42 | 3.18 | 5.230 | 0.185 | 0.285 | 5.426 |
| 09/12/2020 | 11.31 | 2.25 | 5.232 | 0.185 | 0.264 | 5.427 |
| 10/12/2020 | 11.36 | 0.54 | 5.225 | 0.185 | 0.277 | 5.426 |

Therefore, the forecast values of confirmed cases in USA and Qatar shown in Table 9 were computed based on the cubic linear regression model.

## 5. Conclusions

Four important models including classical ARIMA, k^th^ SMA-ARIMA, k^th^ WMA-ARIMA, and k^th^ EWMA-ARIMA have been considered in the prediction of the confirmed, recovery, and death cases of the novel COVID-19 pandemic in the GCC countries, these models have been applied on the daily data from the first case reported in each country until Nov 30, 2020. To compute the best parameter estimates, each model was fitted for 90% of the available data in each country, which is called the in-sample forecast or training data, and the remaining 10% was used for the out-of-sample forecast or testing the model. The AIC was applied to the training data as a criterion method to select the best model. Furthermore, the statistical measure RMSE was utilized for testing data, and the model with the minimum AIC and minimum RMSE was selected. The main finding, in general, is that the two models WMA-ARIMA and EWMA-ARIMA, besides the cubic linear regression model have given better results for in-sample and out-of-sample forecasts than the classical ARIMA models in fitting the confirmed and recovery cases while the death cases haven’t specific models.

### Patient consent

No written consent has been obtained from the patients as there is no patient identifiable data included in this study.

## Data Availability

The data that the findings of this study are openly available.

https://www.who.int/emergencies/diseases/novel-coronavirus-2019/situation-reports

https://sehhty.com/

https://en.wikipedia.org/wiki/COVID-19_pandemic

## Data Availability

The data that the findings of this study are openly available, at the web addresses https://www.who.int/emergencies/diseases/novel-coronavirus-2019/situation-reports. https://sehhty.com/

https://en.wikipedia.org/wiki/COVID-19_pandemic

## APPENDIX

**Table A.1.** ADF unit root test for confirmed, recovery and death cases in each country

| Country | Confirmed cases |  | Recovery cases |  | Death cases |  |
| --- | --- | --- | --- | --- | --- | --- |
|  | Level or difference order | t-test | Level or difference order | t-test | Level or difference order | t-test |
| KSA | 1<sup>st</sup> difference | -8.089*** | 1<sup>st</sup> difference | -11.463** | 1<sup>st</sup> difference | -30.268** |
| UAE | 1<sup>st</sup> difference | -7.921*** | 1<sup>st</sup> difference | -18.152** | 1<sup>st</sup> difference | -3.450*** |
| Kuwait | 1<sup>st</sup> difference | -13.750*** | 1<sup>st</sup> difference | -24.160** | 1<sup>st</sup> difference | -12.771*** |
| Qatar | 1<sup>st</sup> difference | -17.402** | 1<sup>st</sup> difference | -8.596*** | 1<sup>st</sup> difference | -4.987*** |
| Bahrain | 1<sup>st</sup> difference | -17.653*** | 1<sup>st</sup> difference | -12.428*** | 1<sup>st</sup> difference | -3.678*** |
| Oman | 1<sup>st</sup> difference | -7.920*** | 1<sup>st</sup> difference | -18.152** | 1<sup>st</sup> difference | -3.445*** |

Note: \*Significant at the 0.10 level. \*\*Significant at the 0.05 level. \*\*\*Significant at the 0.01 level.

## References

[1] Alandijany, T. A., Faizo, A. A., & Azhar, E. I. (2020). Coronavirus disease of 2019 (COVID-19) in the Gulf Cooperation Council (GCC) countries: Current status and management practices. Journal of infection and public health, 13, 839–842, (doi.org/10.1016/j.jiph.2020.05.020)

[2] Sharif, A. F., Mattout, S. K., & Mitwally, N. A. (2020). Coronavirus disease-19 spread in the Eastern Mediterranean Region, updates and prediction of disease progression in Kingdom of Saudi Arabia, Iran, and Pakistan. International Journal of Health Sciences, 14(5), 32.

[3] Zuo, M., Khosa, S. K., Ahmad, Z., & Almaspoor, Z. (2020). Comparison of COVID-19 pandemic dynamics in Asian countries with statistical modeling. Computational and mathematical methods in medicine, 2020,, 1–16, (doi.org/10.1155/2020/4296806).

[4] Abuhasel, K. A., Khadr, M., & Alquraish, M. M. (2020). Analyzing and forecasting COVID-19 pandemic in the Kingdom of Saudi Arabia using ARIMA and SIR models. Computational intelligence, (doi.org/10.1111/coin.12407).

[5] Ayinde, K., Lukman, A. F., Rauf, R. I., Alabi, O. O., Okon, C. E., & Ayinde, O. E. (2020). Modeling Nigerian Covid-19 cases: A comparative analysis of models and estimators. Chaos, Solitons & Fractals, 138, 109911, (doi.org/10.1016/j.chaos.2020.109911).

[6] Elhassan, T., & Gaafar, A. (2020). Mathematical modeling of the COVID-19 prevalence in Saudi Arabia. medRxiv, (doi.org/10.1101/2020.06.25.20138602).

[7] Singh, S., Sundram, B. M., Rajendran, K., Law, K. B., Aris, T., Ibrahim, H., … & Gill, B. S. (2020). Forecasting daily confirmed COVID-19 cases in Malaysia using ARIMA models. The Journal of Infection in Developing Countries, 14(09), 971–976, (doi.org/10.3855/jidc.13116).

[8] Ding, G., Li, X., Shen, Y., & Fan, J. (2020). Brief Analysis of the ARIMA model on the COVID-19 in Italy. medRxiv, (doi.org/10.1101/2020.04.08.20058636).

[9] Hernandez-Matamoros, A., Fujita, H., Hayashi, T., & Perez-Meana, H. (2020). Forecasting of COVID19 per regions using ARIMA models and polynomial functions. Applied Soft Computing, 96, 106610, (doi.org/10.1016/j.asoc.2020.106610).

[10] Dawoud, I. (2020). Modeling Palestinian COVID-19 Cumulative Confirmed Cases: A Comparative Study. Infectious Disease Modelling, 5, 748–754, (doi.org/10.1016/j.idm.2020.09.001).

[11] Duong, N. Q., Quynh, D. T. N., Loan, C. T. A., & Diem, P. T. H. (2020). Predicting the Pandemic COVID-19 Using ARIMA Model. VNU Journal of Science: Mathematics-Physics, 36(4), 46–57, (doi.org/10.25073/2588-1124/vnumap.4492).

[12] Roy, S., Bhunia, G. S., & Shit, P. K. (2021). Spatial prediction of COVID-19 epidemic using ARIMA techniques in India. Modeling earth systems and environment, 7(2), 1385–1391, (doi.org/10.1007/s40808-020-00890-y).

[13] Verma, P., Khetan, M., Dwivedi, S., & Dixit, S. (2020). Forecasting the covid-19 outbreak: an application of arima and fuzzy time series models, (doi.org/10.21203/rs.3.rs-36585/v1)

[14] Shih, S. H., & Tsokos, C. P. (2008). A weighted moving average process for forecasting. Journal of Modern Applied Statistical Methods, 7(1), 15, (doi.org/10.22237/jmasm/1209615240).

[15] Tsokos, C. P. (2010). K-th moving, weighted and exponential moving average for time series forecasting models. European journal of pure and applied mathematics, 3(3), 406–416.

[16] Akaike, H. (1974). A new look at the statistical model identification. IEEE transactions on automatic control, 19(6), 716–723, (doi.org/10.1109/TAC.1974.1100705).

[17] Hyndman, R. J., & Koehler, A. B. (2006). Another look at measures of forecast accuracy. International journal of forecasting, 22(4), 679–688, (doi.org/10.1016/j.ijforecast.2006.03.001).

[18] Box, G. E. P., Jenkins, G. M., & Reinsel, G. C. (1994). Time series analysis forecasting and control (3rd ed.). New Jersey: Prentice Hall.

